# Parsing the Pediatric Mood Spectrum in U.S. Youth: Transdiagnostic Severity Versus Phenotypic Specificity

**DOI:** 10.64898/2026.10.09.26364902

**Authors:** Kevin G. Stephenson, Phoebe A. Rodda, Carla Mazefsky, Eric A. Youngstrom

## Abstract

**Objective:** Emotion dysregulation and irritability occur across pediatric disorders, complicating distinctions between general symptom burden and specific clinical presentations. Person-centered analyses can look at profiles of scores to identify groups with similar presentations. We examined whether modeling shared and domain-specific symptom variance identified differentiated profiles in youth.

**Method:** Caregivers of 2,187 US youth ages 6–18 completed 87 items assessing emotion dysregulation, depression, mania-related symptoms, sleep-related impairment, and attention and behavioral problems. An ordinal bifactor model estimated one general and 13 specific factors. Latent profile analyses used standardized scores on all 14 factors, with an expanded raw-score analysis as a comparator. We examined caregiver-reported diagnoses and functioning and approximately two-week classification agreement among 166 recontacted caregivers.

**Results:** The general factor accounted for 62% of common variance (ωH=.94), although model fit was mixed (CFI=.884; RMSEA=.058; SRMR=.092). A provisional four-class solution distinguished population-modal (67.4%), internalizing (11.8%), attention/hyperactivity (11.0%), and unusual-thought/elevated-mood (9.8%) profiles. Entropy was .83, but model enumeration and initialization checks did not establish a unique solution. Profiles showed overlapping diagnostic and functional correlates. Recontact agreement was 78.9% (κ=.60). Irritability-specific reliability was limited, constraining interpretation of its weak profile differentiation.

**Conclusions:** General symptom burden coexisted with differentiated residual symptom patterns. These findings support evaluating severity alongside symptom configuration, without equating profiles with diagnoses or weak residual irritability signals with limited clinical relevance. Measurement refinement, model-selection verification, and confirmation of classification-error-adjusted associations are needed before clinical application, but profiles could simplify clinical interpretation of sets of scales.

## Introduction

Pediatric mental health conditions are a substantial US public health concern (AHRQ, 2022; Burnett et al., 2026). Diagnosed conditions increased by 35% between 2016 and 2023, and nearly 20% of US households reported a child needing mental health treatment between June 2023 and September 2024 (Burnett et al., 2026). Unmet need for timely, appropriate treatment (Barnett et al., 2022; Burnett et al., 2026; Wissow et al., 2013) and high psychiatric comorbidity (Ashman et al., 2026; Kessler et al., 2012) underscore the need for parsimonious, well-validated frameworks of pediatric psychopathology that can inform accessible interventions.

### General Factors of Psychopathology and Mood

Research supports a bifactor model of psychopathology encompassing a latent general psychopathology factor (*p*) and narrower construct-specific factors (Caspi et al., 2014; Gluschkoff et al., 2019). The general dimension may capture a tendency toward psychiatric distress across domains, accounting for comorbidity and the co-occurrence of internalizing and externalizing disorders (Caspi et al., 2014; Gluschkoff et al., 2019), consistent with efforts to reorganize psychiatric nosology dimensionally (Kotov et al., 2017). Within mood disorder scales, a general mood factor (*g*) may similarly capture shared variance across manic and depressive symptoms. If *g* reliably accounts for substantial item variance, assessments and interventions addressing this shared dimension could be prioritized. However, its etiology and substantive meaning remain unclear. Emotion dysregulation has been proposed as an observable mechanism through which transdiagnostic affective distress can be measured clinically (Paulus et al., 2021).

### Transdiagnostic Emotion Dysregulation

Emotion dysregulation is a central component of mood disorders, including both bipolar spectrum disorders (BPSDs) and unipolar depression (e.g., major depressive disorder; MDD), as well as common psychiatric comorbidities, such as oppositional defiant disorder, attention-deficit/hyperactivity disorder and generalized anxiety disorder (Aldao et al., 2010; Geise et al., 2014; Khafif et al., 2021). In recent years, emotion dysregulation has emerged as an important transdiagnostic predictor across a wide range of mental health outcomes spanning internalizing, externalizing, thought-disorder, and sleep domains, and has been discussed as a putative explanation for psychiatric comorbidity (Beauchaine & Cicchetti, 2019; Mazefsky et al., 2013; Shaw et al., 2014; White et al., 2014). Emotion dysregulation is often operationalized as an inability to modulate the intensity and duration of emotional experience and responses, such that goal-directed activity is clinically impaired (Mazefsky, Yu, et al., 2018, 2021; Sloan et al., 2017; Thompson, 2019; Weiss et al., 2014). Conceptually, emotion dysregulation spans affective experience, encompassing negative mood and anxiety, and affective control, encompassing reactivity and emotional inflexibility (Mazefsky, Day, et al., 2018). Emotion dysregulation is associated with poor functioning across psychiatric, behavioral, and social domains, and has been studied as a central target within developmental intervention frameworks (Mazefsky, Yu, et al., 2018; Prizant et al., 2003). Transdiagnostic interventions targeting emotion regulation have produced comparable outcomes to disorder-specific treatments in several trials (Barlow et al., 2004; Kolaas et al., 2024; Malmberg et al., 2019). Such approaches may improve accessibility and applicability across clinical presentations and diagnostic groupings (Mazefsky, Day, et al., 2018).

### Emotion Dysregulation & Pediatric Mood Disorders

As in adults, emotion dysregulation has been posited to be an important transdiagnostic factor that confers increased risk for a diverse range of psychopathology outcomes among pediatric populations (McLaughlin et al., 2011). In the context of unipolar depression, chronic emotion dysregulation and maladaptive emotion regulation in childhood and early adolescence have been associated with increased risk of depression and internalizing psychopathology (Defayette et al., 2021; Murray et al., 2025). The prevalence and associated impact of unipolar depressive disorders, such as MDD, among children and adolescents has risen substantially in recent years (Lu et al., 2024). Conversely, interventions aimed at improving emotion regulation skills in youth have demonstrated a protective effect against the development and worsening of depressive symptomatology (Defayette et al., 2021; Murray et al., 2025).

Emotion dysregulation also has been applied to better understand the etiology of pediatric mood disorders, particularly pediatric bipolar disorder (PBD; Khafif et al., 2021). Over the past few decades, PBD has gained increasing recognition as a clinically relevant and well-validated diagnosis (Goldstein et al., 2017). Youth with PBD experience [hypo]mania, characterized by elevated mood, increased energy, or irritability, often alongside periodic depression, characterized by increased negative affect alongside reduced positive affect, and mixed affective states (Birmaher, 2013; A. R. Van Meter et al., 2016). Children and adolescents with PBD often experience emotion regulation difficulties, with lab paradigms documenting issues in the domains of facial emotion recognition and affective language interference (Dickstein et al., 2004; Dickstein & Leibenluft, 2006; Khafif et al., 2021; McClure et al., 2025).

A long-running debate in pediatric mood disorder nosology concerns whether severe, chronic emotion dysregulation (labeled in some disciplines as irritability or mood lability) irritability constitutes a developmental phenotype of bipolar disorder in youth. Through the 1990s and 2000s, an influential view held that pediatric bipolar disorder presents not with the discrete manic episodes seen in adults but with chronic, non-episodic irritability and mood lability (Leibenluft et al., 2024) — a formulation that, because it overlaps substantially with so-called externalizing disorders (attention-deficit/hyperactivity disorder [ADHD], oppositional defiant disorder, and conduct disorder), coincided with a marked rise in pediatric bipolar diagnosis rates concentrated in the United States (Sun et al., 2026). Subsequent longitudinal and cross-national work reframed the debate: chronic irritability was found to predict adult depressive and anxiety disorders rather than mania (Leibenluft et al., 2024), symptom trajectories separating juvenile bipolar disorder from ADHD were driven by elevated mood and decreased need for sleep rather than irritability, which followed parallel trajectories in both groups (Luckenbaugh et al., 2009). DSM-5 attempted to resolve this by introducing disruptive mood dysregulation disorder (DMDD) — notably classified among the depressive disorders, not the bipolar-spectrum or disruptive-behavior disorders — and by explicitly restricting “bipolar disorder” to episodic presentations (American Psychiatric Association (APA), 2022).

The nosology remains unsettled internationally, however: ICD-11 did not adopt a parallel freestanding diagnosis, instead treating chronic irritability as more consistent with a mood/emotional-dysregulation specifier on oppositional defiant disorder than with its own disorder category (Evans et al., 2017). The 2017 International Society for Bipolar Disorders Task Force concluded the core diagnostic question with regard to bipolar disorder is nonetheless largely resolved: chronic irritability, regardless of severity, is not by itself sufficient for a bipolar diagnosis, though it can count toward manic/hypomanic criteria when it emerges or intensifies alongside other Criterion B symptoms (Goldstein et al., 2017). Whether an irritability-specific signal, once separated from general mood-symptom severity, shows meaningful incremental or discriminant validity for any specific mood or behavioral phenotype remains an open empirical question this study’s bifactor approach is well positioned to address directly.

Recent research has also focused on exploring the interplay between emotion regulation and sleep in youth (Sesso et al., 2024). Sleep disturbances are characteristic of or associated with many pediatric psychopathologies, bridging physiologic and psychologic domains. Youth with mood disorders may be especially prone to sleep disturbances (American Psychiatric Association [APA], 2022), and decreased need for sleep appears to be of particular importance in identifying and discriminating cases of PBD (Baroni et al., 2012). When studied as an independent construct, sleep disturbances have been associated with increased affective instability, internalizing problems, and mood disorder treatment resistance in youth (Clarke & Harvey, 2012; Kansagra, 2020; Sesso et al., 2024). Sleep problems are associated with functional changes in brain regions associated with emotion regulation and integration, and the deleterious effects of reduced sleep during adolescence may be of particular concern (Walker, 2010). While the mechanisms underlying the observed association between emotion dysregulation and impaired sleep are not yet well-defined, it is likely that the relationship between sleep and emotion regulation is bidirectional (Avvenuti et al., 2021; Menz et al., 2013; Sesso et al., 2024; Wang et al., 2024).

### The Present Study

We used bifactor confirmatory factor analysis to assess whether a range of emotional, behavioral, and sleep characteristics (i.e., transdiagnostic emotion dysregulation, ADHD and externalizing characteristics, mania, depression, and sleep-related impairment) reflect both a general emotional-behavioral factor (*g*) and distinct domain-specific factors. We hypothesized that latent profile analysis incorporating bifactor-purified, instrument-specific scores would recover classes differing in residual symptom patterns as well as overall severity. Evidence-based assessment guidelines recommend scales to differentiate unipolar depression and bipolar disorder from other conditions (Becker-Haimes et al., 2020; Dougherty et al., 2018; Hankin & Cohen, 2020; Johnson et al., 2018; E. A. Youngstrom, 2026) and evaluation of sleep in comprehensive mood assessment (E. A. Youngstrom, 2026). Whether these measures retain sufficient specific variance to identify nonuniform profiles beyond general emotion dysregulation remains an empirical question.

## Method

### Sample

The sample consisted of *N*=2,252 caregivers of youth ages 6-18, recruited through YouGov and representative of the general US population; data collection occurred in fall of 2025. Analyses reported here use the *N*=2,187 caregivers with complete data across all 87 items of the full symptom battery. Participants all provided informed consent and were recruited and surveyed through YouGov, an international online survey company, as part of a larger initiative to assess the psychometric properties of 15 caregiver- and youth-report mental, behavioral, and physical health assessments. All study procedures were approved by the Institutional Review Board at MASKED FOR PEER REVIEW, and data were collected between July 11, 2025, and March 7, 2026. The design and analyses were preregistered (E. Youngstrom et al., 2025). Table 1 reports participant characteristics.

**Table 1.** Participant Characteristics.

Participant Characteristics
| Characteristic | N=2,187<sup>1</sup> |
| --- | --- |
| **Child Age** |  |
| 6 years old | 159 (7.3%) |
| 7 years old | 178 (8.1%) |
| 8 years old | 173 (7.9%) |
| 9 years old | 153 (7.0%) |
| 10 years old | 176 (8.0%) |
| 11 years old | 137 (6.3%) |
| 12 years old | 211 (9.6%) |
| 13 years old | 210 (9.6%) |
| 14 years old | 217 (9.9%) |
| 15 years old | 216 (9.9%) |
| 16 years old | 216 (9.9%) |
| 17 years old | 121 (5.5%) |
| 18 years old | 20 (0.9%) |
| **Child Biological Sex** |  |
| Male | 1,149 (52.5%) |
| Female | 1,038 (47.5%) |
| Intersex/Other | 0 (0.0%) |
| **Caregiver Ethnicity** |  |
| Hispanic or Latino | 347 (15.9%) |
| Not Hispanic or Latino | 1,765 (80.7%) |
| Other | 36 (1.6%) |
| Prefer not to say | 39 (1.8%) |
| **Caregiver Race** |  |
| White | 1,731 (79.1%) |
| Black or African American | 213 (9.7%) |
| Asian | 102 (4.7%) |
| Native American / Alaska Native | 81 (3.7%) |
| Native Hawaiian / Pacific Islander | 11 (0.5%) |
| Other | 119 (5.4%) |
| Prefer not to say | 34 (1.6%) |
| **Child Diagnoses (Caregiver-Reported)** |  |
| Depression | 183 (8.4%) |
| Anxiety | 345 (15.8%) |
| ADHD | 467 (21.4%) |
| Bipolar Disorder | 25 (1.1%) |
| Autism Spectrum | 174 (8.0%) |
| Disruptive Behavior | 74 (3.4%) |
| Sleep Disorder | 94 (4.3%) |
| Learning Disability | 170 (7.8%) |
| None | 1,307 (59.8%) |
| **Caregiver Education** |  |
| No high school diploma | 51 (2.3%) |

| **Characteristic** | **N=2,187<sup>1</sup>** |
| --- | --- |
| High school graduate | 382 (17.5%) |
| Some college | 446 (20.4%) |
| 2-year degree | 261 (11.9%) |
| 4-year degree | 609 (27.8%) |
| Post-grad | 438 (20.0%) |
| **Family Income** |  |
| Less than \$10,000 | 92 (4.2%) |
| \$10,000 - \$19,999 | 88 (4.0%) |
| \$20,000 - \$29,999 | 143 (6.5%) |
| \$30,000 - \$39,999 | 132 (6.0%) |
| \$40,000 - \$49,999 | 131 (6.0%) |
| \$50,000 - \$59,999 | 164 (7.5%) |
| \$60,000 - \$69,999 | 152 (7.0%) |
| \$70,000 - \$79,999 | 194 (8.9%) |
| \$80,000 - \$99,999 | 200 (9.1%) |
| \$100,000 - \$119,999 | 202 (9.2%) |
| \$120,000 - \$149,999 | 240 (11.0%) |
| \$150,000 - \$199,999 | 184 (8.4%) |
| \$200,000 - \$249,999 | 77 (3.5%) |
| \$250,000 - \$349,999 | 49 (2.2%) |
| \$350,000 - \$499,999 | 24 (1.1%) |
| \$500,000 or more | 10 (0.5%) |
| Prefer not to say | 105 (4.8%) |
| **Geographic Region** |  |
| Northeast | 415 (19.0%) |
| Midwest | 484 (22.1%) |
| South | 825 (37.7%) |
| West | 463 (21.2%) |
| **Residence Type** |  |
| Big city | 384 (17.6%) |
| Smaller city | 339 (15.5%) |
| Suburban area | 895 (40.9%) |
| Small town | 255 (11.7%) |
| Rural area | 314 (14.4%) |

<sup>1</sup>n (%)

### Measures

#### Caregiver Report Child Mania Rating Scale (CMRS-P)

The CMRS-P is a 21-item caregiver-report screening measure that is commonly used to measure DSM-IV-TR PBD symptoms (Pavuluri et al., 2006). Caregivers rate youth mood and behavior over the past month using a 4-point Likert scale (0: Never/Rare, 1: Sometimes, 2: Often, 3: Very Often), with higher scores being indicative of greater manic symptom severity.

While the CMRS-P has traditionally been treated as having a single total score (Pavuluri et al., 2006), factor analyses (McClellan et al., in press) support the existence of four factors: Irritable Mood, Elevated Mood, Strange Thoughts & Behaviors, and Increased Activity.

#### Very Quick Inventory of Depressive Symptomatology (VQIDS-5)

The VQIDS-5 is a 5-item short form derived from the QIDS-16 that is used to measure core depressive symptoms, including sad mood, poor self-outlook, loss of interest or involvement, fatigue, and psychomotor slowing (De La Garza et al., 2017; Rush et al., 2026). Each item uses a Likert-type scale from 0 to 3, creating a possible range from 0 to 15, with high scores indicating more depressive symptom severity. The self-report VQIDS-5 (VQIDS-5-SR) demonstrates a single factor structure and adequate internal consistency, sensitivity to change, reliability, and relationship with overall depression severity (De La Garza et al., 2017; Rush et al., 2026). Caregivers completed the VQIDS-P-A, an unpublished caregiver-report version of the VQIDS-5-SR created by authors of the original measure.

#### PROMIS Pediatric Sleep-Related Impairment Item Bank (PROMIS-SRI)

The PROMIS-SRI is a 13-item bank assessing daytime sleepiness, sleep offset, and the impact of sleepiness on cognitive functioning, affect and behaviors, and daily activities (Forrest et al., 2018). Items are scored on a 5-point Likert scale (1: Never, 2: Almost Never, 3: Sometimes, 4: Almost Always, 5: Always). The PROMIS-SRI has demonstrated content and structural validity, excellent reliability and precision across severity, and strong internal consistency (Becker et al., 2026; Forrest et al., 2018). Caregivers completed the PROMIS-SRI Parent Proxy v1.0, reporting their child’s symptoms over the past week.

#### Emotion Dysregulation Inventory (EDI)

The EDI is a 24-item measure of pediatric emotion dysregulation (Mazefsky, Day, et al., 2018; Mazefsky, Yu, et al., 2018). While the EDI was originally developed to assess emotion dysregulation in youth with ASD, it is broadly applicable as an efficient and precise measure of pediatric emotion dysregulation in both clinical and community samples (Mazefsky, Yu, et al., 2021). The EDI is a valid, reliable, and change-sensitive measure with two subscales that are correlated but scored independently: reactivity, including both initial emotional reactivity and difficulty down-regulating negative emotion, and dysphoria, capturing lower arousal and negative valence characterized by general unease (Mazefsky, Yu, et al., 2018, 2021). Caregivers completed the 7-item short form of the reactivity subscale (EDI-R-SF) and the 6-item dysphoria subscale (EDI-D), rating youth symptoms over the past week using a Likert-type scale ranging from 0 (Not At All) to 4 (Very Severe).

#### Vanderbilt ADHD Diagnostic Rating Scale (VADRS)

The VADRS assesses inattentive and hyperactive-impulsive ADHD symptoms across two 9-item subscales covering all DSM symptom criteria (Wolraich et al., 2003), typically completed by a parent or teacher. Caregivers completed the parent-report VADRS, rating each item as Never (0), Occasionally (1), Often (2), or Very Often (3). Vanderbilt-Inattention and Vanderbilt-Hyperactivity/Impulsivity served as two further core specific factors within the bifactor model. The VADRS also includes an 8-item performance/impairment section spanning academic (4 items) and classroom-behavioral/social (4 items) functioning; these were scored separately as continuous validators of academic and social impairment rather than entered into the bifactor model, since they index functional impact rather than symptom severity.

#### Pediatric Symptom Checklist-17 (PSC-17)

The PSC-17 assesses internalizing, externalizing, and attention problems across three subscales (Gardner et al., 1999). Widely used in screening contexts to identify youth who would benefit from further evaluation, the measure is well suited for measurement-based-care applications given its brevity and subscale specificity. Caregivers completed the parent-report PSC-17, rating each item as Never (0), Sometimes (1), or Often (2). The PSC-17’s three subscales served as core specific factors within the bifactor model reported throughout this paper.

### Impairment and Functioning

#### Nationwide Gauge of Emotional Severity (N-Gauge)

The N-Gauge is a single-item, caregiver-rated measure of youth global emotional functioning on a 6-point scale (0–5), analogous to the Clinical/Patient Global Impressions severity scale (McClellan et al., 2025).

#### Nationwide Quality of Life Scale (NQLS)

The NQLS is a 7-item scale assessing satisfaction across seven life domains: friends, family, grades, school in general, health, life, and anticipated future. Originally developed for youth self-report (Covarrubias & Fristad, 2025), the NQLS has since been evaluated for parent-report within a large nationally representative community sample (P-NQLS; Liu et al., 2026).

Caregivers completed the P-NQLS, rating each item on a five-point Likert scale (0=Very Unsatisfied to 4=Very Satisfied); total scores range 0–28, where higher scores indicate greater parent-perceived youth quality of life.

#### Family Index of Risk for Mood (FIRM)

The FIRM is a caregiver-report measure assessing a youth’s family-history risk for mood disorders based on relatives’ history of five concerns — suicide, alcohol/drug problems, psychiatric hospitalization, depression, and manic/bipolar episodes — each rated across four relative categories (grandparents, parents, aunts/uncles, and siblings), yielding 20 checklist items, plus two further items assessing the responding parent’s own lifetime history of bipolar disorder and depression (Algorta et al., 2013). Each item is scored as endorsed (1) or not endorsed (0) and summed into a 22-item total. In our sample, FIRM-22 total scores ranged from 0 to 19 (*M*=3.20; *SD*=3.30).

### Demographics and Prior Services and Diagnoses

In addition, parents completed demographic variables, including youth sex as a binary, youth age, and family income – selected as a multiple choice from a set of income bands. Parents also reported whether the youth had previously been diagnosed with mental or behavioral health conditions, and whether they had taken medication, had therapy, or used supplements or alternative treatments to address mental/behavioral issues.

### Data Analysis

#### Mood Dimensionality Analysis

To evaluate the extent to which mood symptoms reflect a general mood factor versus distinct domain-specific factors, we specified a bifactor confirmatory factor analysis (CFA) using individual items from each measure. Structural validity and score utility were evaluated using omega hierarchical (ω_H_ for general factor; ω_HS_ for specific factors), explained common variance (ECV for the general factor; ECV_SS_ for the specific scales) and construct replicability (*H*). Following established bifactor guidelines (Gignac & Kretzschmar, 2017; Rodriguez et al., 2016), benchmarks of ω_H_ ≥ .80, ω_HS_ ≥ .20, ECV_SS_ ≥ .30, and *H* ≥ .70 were applied to evaluate general factor reliability, subscale utility beyond *g*, variance attributable to the specific factor, and latent trait replicability, respectively.

### Measurement model

A confirmatory bifactor model was specified across all 87 items from eight instruments (CMRS-P, VQIDS, PROMIS Pediatric Sleep-Related Impairment, EDI-Reactivity, EDI-Dysphoria, PSC-17 Internalizing/Externalizing/Attention subscales, and the Vanderbilt ADHD Diagnostic Parent Rating Scale Inattention and Hyperactivity/Impulsivity subscales), estimated with a diagonally weighted least squares estimator (WLSMV), the recommended estimator for confirmatory factor models with ordinal/Likert-type indicators (Flora & Curran, 2004). The model specified one general factor (g, reflecting transdiagnostic mood/behavioral severity) loading on all 87 items, and 13 orthogonal specific factors, each loading only on its own instrument’s or subscale’s items, corresponding to four factors underlying the CMRS--Irritable Mood, Elevated Mood, Strange Thoughts, and Increased Activity (McClellan et al., under review); VQIDS depression (Rush et al., 2022; Algorta et al., under review), EDI-Reactivity and- Dysphoria (Mazefsky, Yu, et al., 2018); PROMIS-Sleep (Becker et al., 2026; Forrest et al., 2018), PSC-Internalizing, Externalizing, and Attention (Gardner et al., 1999); Vanderbilt-Inattention, and Hyperactivity/Impulsivity (Wolraich et al., 2003; Evans et al., under review). Supplemental Figures S1 & S2 present the measurement model and indicators.

This bifactor parameterization follows recommended practice for multi-instrument batteries expected to share substantial general-severity variance (Reise, 2012): removing that shared variance from each instrument’s specific factor is a precondition for any downstream profile analysis to recover differentiated phenotypic content rather than a severity gradient (see Latent profile analysis, below). PSC-Attention items were retained as their own specific factor rather than merged with the conceptually overlapping Vanderbilt ADHD subscales: although PSC-Attention correlated strongly with Vanderbilt Inattention/Hyperactivity at the raw-score level (*r*=.74–.77), that overlap was shown to be almost entirely attributable to shared general-factor variance — the purified, g-orthogonalized specific-factor correlation dropped to *r*=.28–.35 — and PSC-Attention’s specific factor showed a distinct external correlational signature (most notably a robust negative association with the CMRS-Irritable Mood specific factor not shared by the Vanderbilt factors), indicating it captured non-redundant information.

### Latent profile analysis

Factor scores for the general factor and all 13 specific factors were extracted from the bifactor model and standardized. Latent profile analysis was conducted on this 14-indicator, fully bifactor-purified set using Gaussian finite mixture models with equal variances and covariances across classes (the EEE parameterization; Scrucca et al., 2016), for 1 to 7 classes. Model selection did not rely on Bayesian Information Criterion (BIC) alone: BIC improved monotonically without a clear inflection point through the tested range, a pattern found in preliminary analyses to reflect model instability rather than genuine additional structure. The retained solution was instead selected using a combination of BIC, entropy, bootstrapped likelihood ratio tests, and a multi-seed stability check (5 random restarts per candidate solution), retaining only solutions for which independent restarts converged to statistically indistinguishable partitions (adjusted Rand index ≥ .65 across restarts; Hubert & Arabie, 1985). A 2-class solution (severity gradient only) was found to be essentially perfectly stable (adjusted Rand index=1.0 across restarts) and is reported as a robustness floor; a 4-class solution showed acceptable (though not perfect) stability (adjusted Rand index=.69–.91) and was retained as the primary solution because its four classes nested almost exactly within the stable 2-class partition (98–100% concordance) and were independently validated against external criteria (below).

### Recontact-sample classification stability

A pre-registered 2-week parent-proxy recontact subsample (N=171 passing a strict target-identity match requiring either a single-child household or unchanged child sex with no implausible age/grade/height drift; N=166 with complete item data at both timepoints) provided an independent, real-world test of classification stability distinct from the internal multi-seed check above. Time-2 item responses were scored through the fixed (not refit) bifactor model to place Time-2 factor scores on the same metric as the retained solution, and each case was then classified into the fixed four-class solution using its original class definitions. Time-1 and Time-2 class assignments for the same individuals were compared via raw percent agreement, Cohen’s kappa, and Gwet’s AC1 and AC2 (Cohen, 1960; Gwet, 2008, 2014), the latter included alongside kappa because kappa can understate agreement under the kind of skewed class-prevalence distribution present here (Normative ≈69%).

### Classification-uncertainty correction

Because latent class assignment is probabilistic rather than certain, all associations between class membership and external validators were estimated using the Bolck-Croon-Hagenaars (BCH) correction (Bolck et al., 2004), which weights a pseudo-class-expanded dataset by the inverse of the model’s own classification-error matrix, yielding an association estimate corrected for classification uncertainty rather than one conditioned on error-prone modal class assignment. Outcomes were modeled as weighted least-squares regressions (equivalent to a linear probability model for binary criteria) with cluster-robust standard errors (clustered on the original case, since each case contributes four weighted pseudo-observations), estimated by direct weighted-least-squares matrix solution rather than through lm()/glm(), both of which reject the negative weights that the BCH correction can produce under imperfect class separation.

### Criterion validation

Latent classes were validated against three families of external criteria, all BCH-corrected and referenced to the Normative class: (1) eight caregiver-reported categorical diagnoses (depression, anxiety, ADHD, bipolar disorder, autism, aggression/disruptive behavior, sleep disorder, learning disability) and three mental-health service-use indicators (medication, over-the-counter/supplement use, therapy); (2) six continuous functional/severity validators (quality of life, global clinical severity, family impact, suicidality risk, and academic/social impairment); and (3) demographic composition (child age, sex, family income, and parent-reported pubertal development), analyzed separately from the clinical criteria given its distinct interpretive purpose. For each outcome, BCH-weighted contrasts with the Normative class were estimated by weighted least-squares regression with cluster-robust standard errors.

#### Incremental Validity of Domain-Specific and Transdiagnostic Mood Symptoms

To evaluate the incremental validity of transdiagnostic mood symptoms, we estimated multiple logistic regression models examining caregiver-reported clinical outcomes. Continuous predictors were standardized prior to analysis such that odds ratios represent the change in odds associated with a 1-*SD* increase in each predictor. Each model simultaneously included both EDI subscales (EDI-Reactivity and EDI-Dysphoria) and the relevant domain-specific measure: PROMIS Sleep for sleep disorder, VQIDS for depression, CMRS-P 21 for bipolar disorder, and CMRS-P Irritable Mood for aggression.

## Results

### Mood Dimensionality Analysis

The final bifactor model — a single general factor (*g*) spanning all 87 items across eight instruments, plus 13 orthogonal specific factors — was justified over a simpler correlated-factors model because the eight source instruments were expected, and confirmed, to share substantial general-severity variance that would otherwise contaminate each instrument’s specific-factor scores and bias any downstream profile solution toward a severity gradient rather than differentiated phenotypic content (see Method: Latent profile analysis). The bifactor model fit the data adequately for a large, 87-item, ordinal, multi-instrument pool (CFI=.884, TLI=.878, RMSEA=.058, SRMR=.092). These values fall short of the CFI ≥ .95 / RMSEA ≤ .06 benchmarks conventionally cited for continuous-indicator models (Hu & Bentler, 2009), but simulation work on WLSMV-estimated categorical models shows those benchmarks are systematically too strict for ordinal data and routinely reject correctly specified models, particularly as the number of items and response categories grows (Xia & Yang, 2019). By the more calibrated standards appropriate to this estimator and item pool, present fit is adequate. All 87 items loaded significantly on *g* (standardized loadings .35–.79) and on their respective specific factors (.09–.75), consistent with the bifactor model’s core assumption that every item carries both transdiagnostic severity variance and construct-specific variance worth separating before profile extraction. The general factor was also reliably and precisely measured (ωH=.94, ECV=.62, *H*=.99, all comfortably clearing benchmark; Table 2). Specific-factor quality varied substantially: PROMIS-Sleep was the strongest (ωHS=.60, ECVSS=.61, H=.92) and PSC-Attention was the only specific factor to clear none of the three benchmarks (ωHS=.14, ECVSS=.19, H=.43) — retained as its own factor regardless, as an explicit exception (see Table 2 note and Method: Measurement model). CMRS-Irritable Mood and EDI-Dysphoria were tied for the next-weakest (ωHS=.23 each), both still clearing the ωHS ≥ .20 floor. Figure 1 shows the bifactor item loading profile.

**Figure 1.**
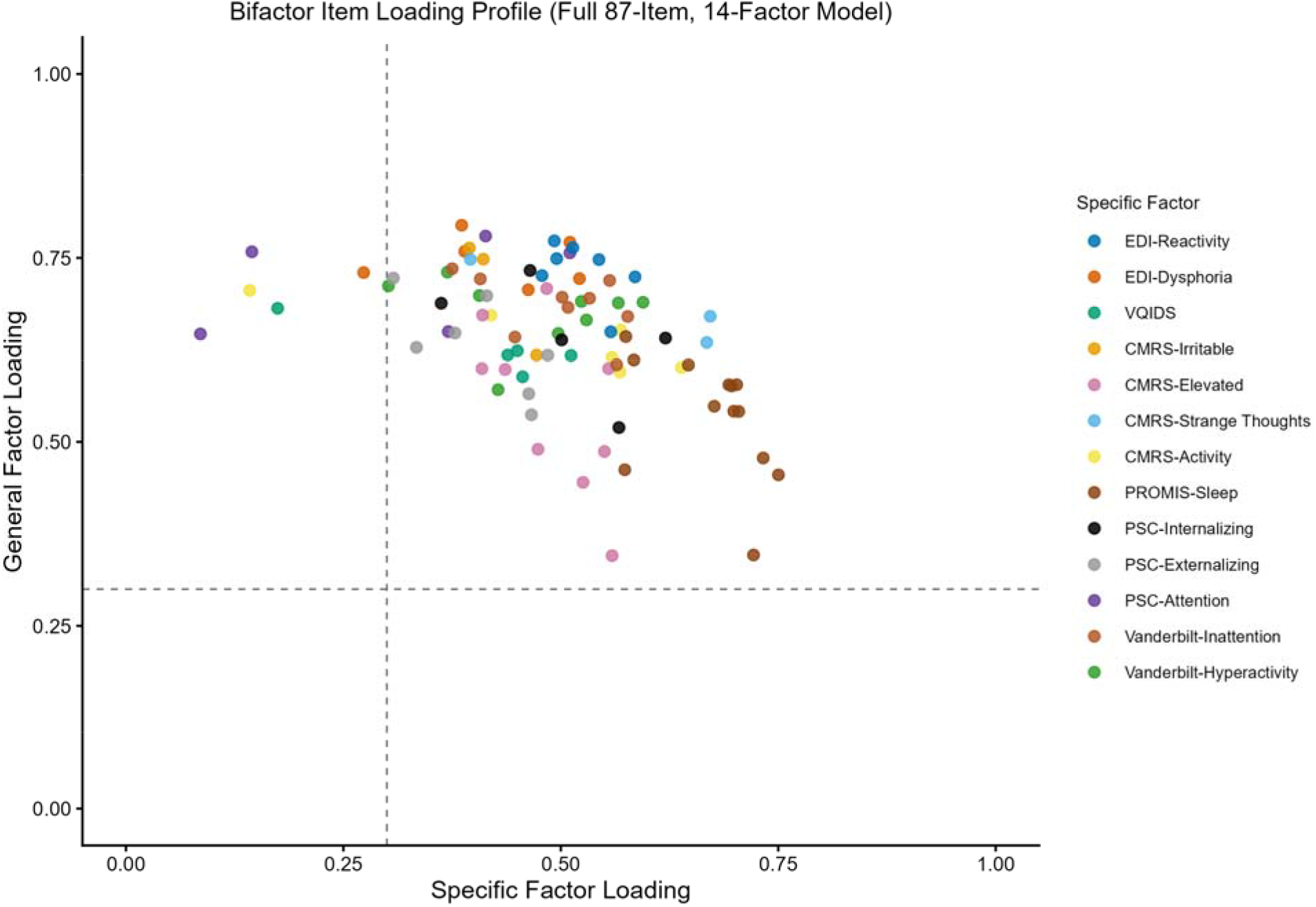
Bifactor Item Loading Profile (Full 87-Item, 14-Factor Model)

**Table 2.**
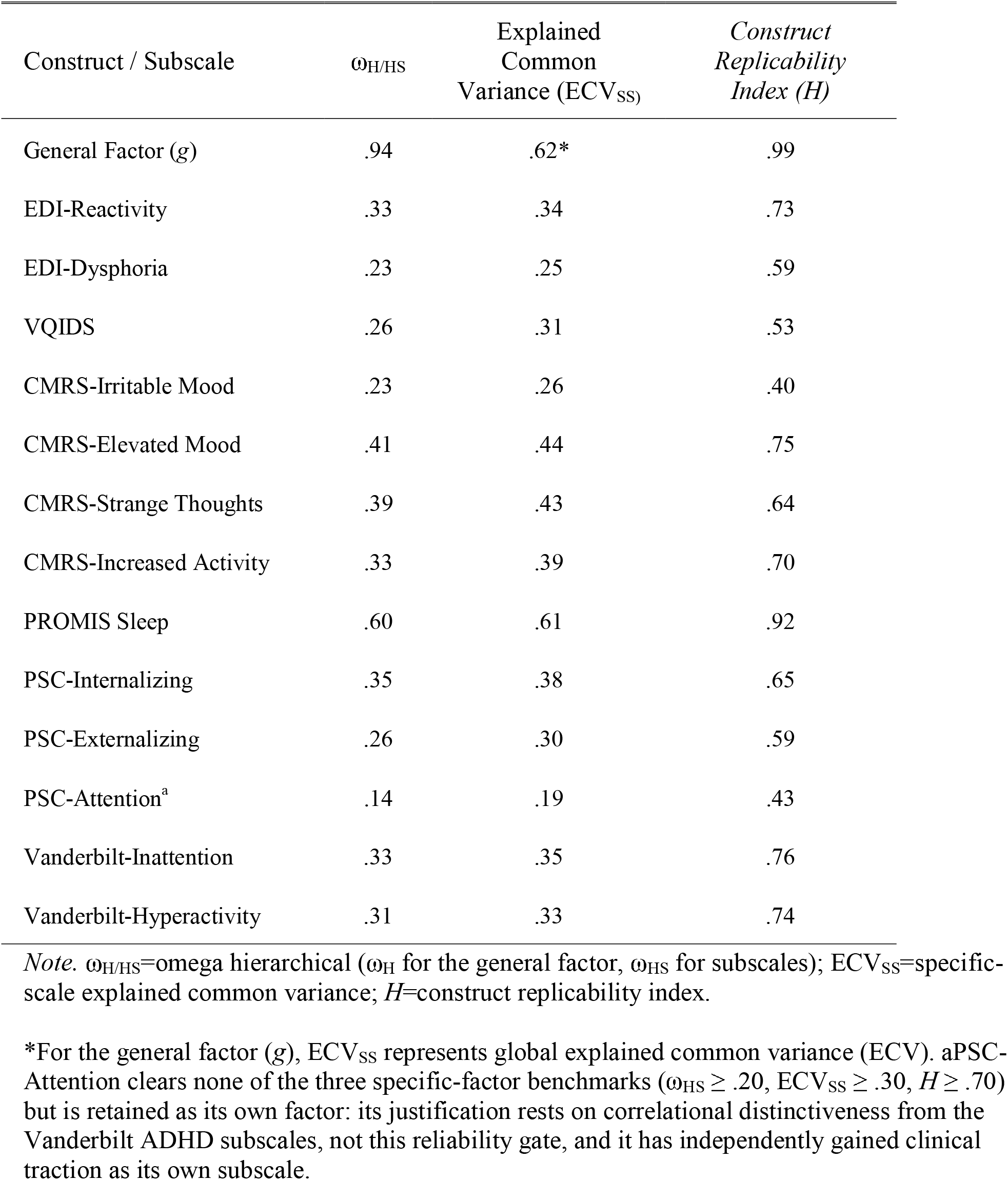
Variance attributable to general and specific factors underlying the battery.

| Construct / Subscale | $\omega_{H/HS}$ | Explained<br>Common<br>Variance ($ECV_{SS}$) | *Construct*<br>*Replicability*<br>*Index (H)* |
| --- | --- | --- | --- |
| General Factor (*g*) | .94 | .62* | .99 |
| EDI-Reactivity | .33 | .34 | .73 |
| EDI-Dysphoria | .23 | .25 | .59 |
| VQIDS | .26 | .31 | .53 |
| CMRS-Irritable Mood | .23 | .26 | .40 |
| CMRS-Elevated Mood | .41 | .44 | .75 |
| CMRS-Strange Thoughts | .39 | .43 | .64 |
| CMRS-Increased Activity | .33 | .39 | .70 |
| PROMIS Sleep | .60 | .61 | .92 |
| PSC-Internalizing | .35 | .38 | .65 |
| PSC-Externalizing | .26 | .30 | .59 |
| PSC-Attention<sup>a</sup> | .14 | .19 | .43 |
| Vanderbilt-Inattention | .33 | .35 | .76 |
| Vanderbilt-Hyperactivity | .31 | .33 | .74 |

*Note.* $\omega_{H/HS}$ =omega hierarchical ( $\omega_H$ for the general factor, $\omega_{HS}$ for subscales); $ECV_{SS}$ =specific-scale explained common variance; $H$ =construct replicability index.
\*For the general factor (*g*), $ECV_{SS}$ represents global explained common variance (ECV). aPSC-Attention clears none of the three specific-factor benchmarks ( $\omega_{HS} \geq .20$ , $ECV_{SS} \geq .30$ , $H \geq .70$ ) but is retained as its own factor: its justification rests on correlational distinctiveness from the Vanderbilt ADHD subscales, not this reliability gate, and it has independently gained clinical traction as its own subscale.

### Latent Profile Analysis

The four-class solution was retained as the best-supported trade-off across information criteria, nontrivial class sizes, profile interpretability, nesting within the stable two-class severity structure, and theoretically coherent external validation. Although higher-class solutions improved some information criteria, they did not provide a comparably stable or interpretable solution. The four classes comprised a Normative (low symptom severity) class (*n*=1,473, 67.4%), a Depressive/Internalizing class (*n*=259, 11.8%; elevated EDI-Dysphoria, VQIDS, and PSC-Internalizing; below-average Vanderbilt-Hyperactivity), a Reactive/ADHD class (*n*=240, 11.0%; elevated PSC-Attention and both Vanderbilt ADHD subscales; below-average EDI-Dysphoria and PROMIS-Sleep), and a Thought/Mania class (*n*=215, 9.8%; a pronounced, relatively isolated elevation on CMRS-Strange Thoughts, *z*=2.65, alongside more modest elevation on CMRS-Elevated Mood). Profile shapes crossed rather than ran parallel across indicators (Figure 2, top panel), indicating differentiated phenotypic content rather than a single severity gradient. The general factor itself did not fully capture this distinction, ranking only eighth of fourteen indicators in its own between-class discriminating power (Figure 2, bottom panel). Classification precision for the retained four-class solution was acceptable (entropy=.83; mean posterior probabilities=.83–.97).

**Figure 2.**
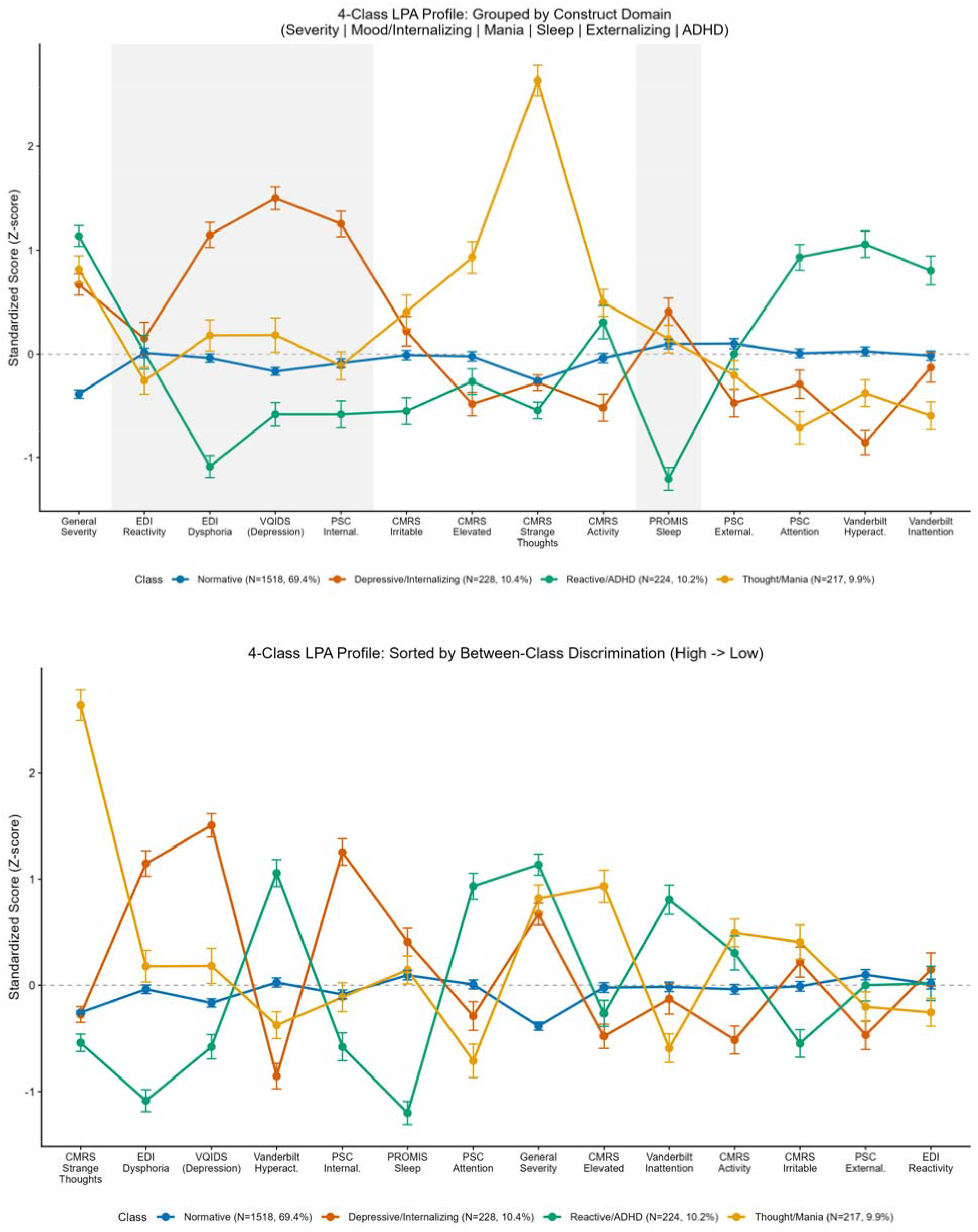
Class Profile Plots, Grouped by Construct Domain (top panel) and Sorted by Between-Class Discrimination (bottom panel)

The retained four-class solution also showed good real-world classification stability. Among 166 recontacted parents whose retest responses passed the protocol’s strict same-child match and had complete item data at both timepoints, raw agreement between Time-1 and Time-2 class assignment was 78.9% (Supplemental Table S5), with Cohen’s kappa=.600 (95% CI [.487, .713]) and Gwet’s AC1=AC2=.744 (95% CI [.666, .823]) — Gwet’s AC1 running appreciably higher than kappa here, consistent with kappa’s known tendency to understate agreement when one category (Normative, 67% of the full sample) dominates. Recurrence was highest for Normative (88.8% reclassified the same way) and lower, though still well above chance, for the three symptomatic classes (Depressive/Internalizing 53.3%, Reactive/ADHD 72.7%, Thought/Mania 57.1%); misclassifications were concentrated as movement toward or from Normative rather than confusion among the three symptomatic classes themselves (no Reactive/ADHD case was reclassified Depressive/Internalizing or vice versa). This complements the internal multi-seed stability check (which asks whether refitting reproduces the solution) with direct evidence that a genuine second administration on the same people, roughly two weeks apart, recovers materially the same classification.

### Criterion Validation

Criterion validation showed a consistent triple dissociation (Supplemental Table S3). ADHD, autism, aggression, and learning-disability diagnoses, along with academic and social impairment, were most pronounced in the Reactive/ADHD class (e.g., ADHD +49.8 percentage points over the Normative base rate, *p* < .00005). Depression and anxiety diagnoses, therapy utilization, and the highest global severity (N-Gauge) were most pronounced in the Depressive/Internalizing class (depression +25.4 percentage points, anxiety +32.7 percentage points, both p < .0001; therapy +46.0 percentage points, *p* < .00005). Bipolar-disorder and sleep-disorder diagnoses were most strongly associated with the Thought/Mania class (bipolar +5.6 percentage points on a 0.1% base rate; sleep disorder +6.2 percentage points, *p*=.003). All three symptomatic classes showed elevated suicidality risk and reduced quality of life relative to Normative (all *p* < .0001), supporting general clinical validity alongside the phenotype-specific pattern.

The classes also differed demographically in a pattern consistent with known developmental epidemiology. Depressive/Internalizing was older (+1.9 years), less likely to be male (–12.2 percentage points), and more pubertally advanced (+2.6 points) than Normative; Reactive/ADHD was younger (–2.0 years), more likely to be male (+22.3 percentage points), and less pubertally advanced (–3.1 points). Thought/Mania was modestly older (+0.7 years) and more pubertally advanced (+1.2 points) than Normative, but did not differ in sex composition. All three symptomatic classes showed modest family-income disadvantage relative to Normative (–0.8 to –1.0 income brackets, all *p* < .05).

### Incremental Validity of Domain-Specific and Transdiagnostic Mood Symptoms

In multiple logistic regression models, domain-specific measures were significantly associated with their corresponding caregiver-reported clinical outcomes (OR range, 1.41–2.11; all *p*_FDR_ ≤ .003). Transdiagnostic EDI subscales demonstrated selective incremental associations with clinical outcomes. EDI-Dysphoria was independently associated with depression (OR=1.85; 95% CI [1.50, 2.28]; *p*_FDR_ < .001), but it was not significantly associated with sleep-related disorders (OR=1.30; 95% CI [1.00, 1.68]; *p*_FDR_=.099), aggression (*p*_FDR_=.795), or bipolar disorder (*p*_FDR_=.896). EDI-Reactivity showed a different pattern: it was independently associated with aggression (OR=1.62; 95% CI [1.21, 2.16]; *p*_FDR_=.004) and negatively associated with a caregiver-reported diagnosis of depression (OR=0.72; 95% CI [0.57, 0.88]; *p*_FDR_=.006), but was not significantly associated with sleep-related disorders (*p*_FDR_=.591) or bipolar disorder (*p*_FDR_=.591). Neither EDI subscale was independently associated with bipolar disorder, beyond CMRS-P 21 (*p*_FDR_ ≥ .591 for both; Figure 3).

**Figure 3.**
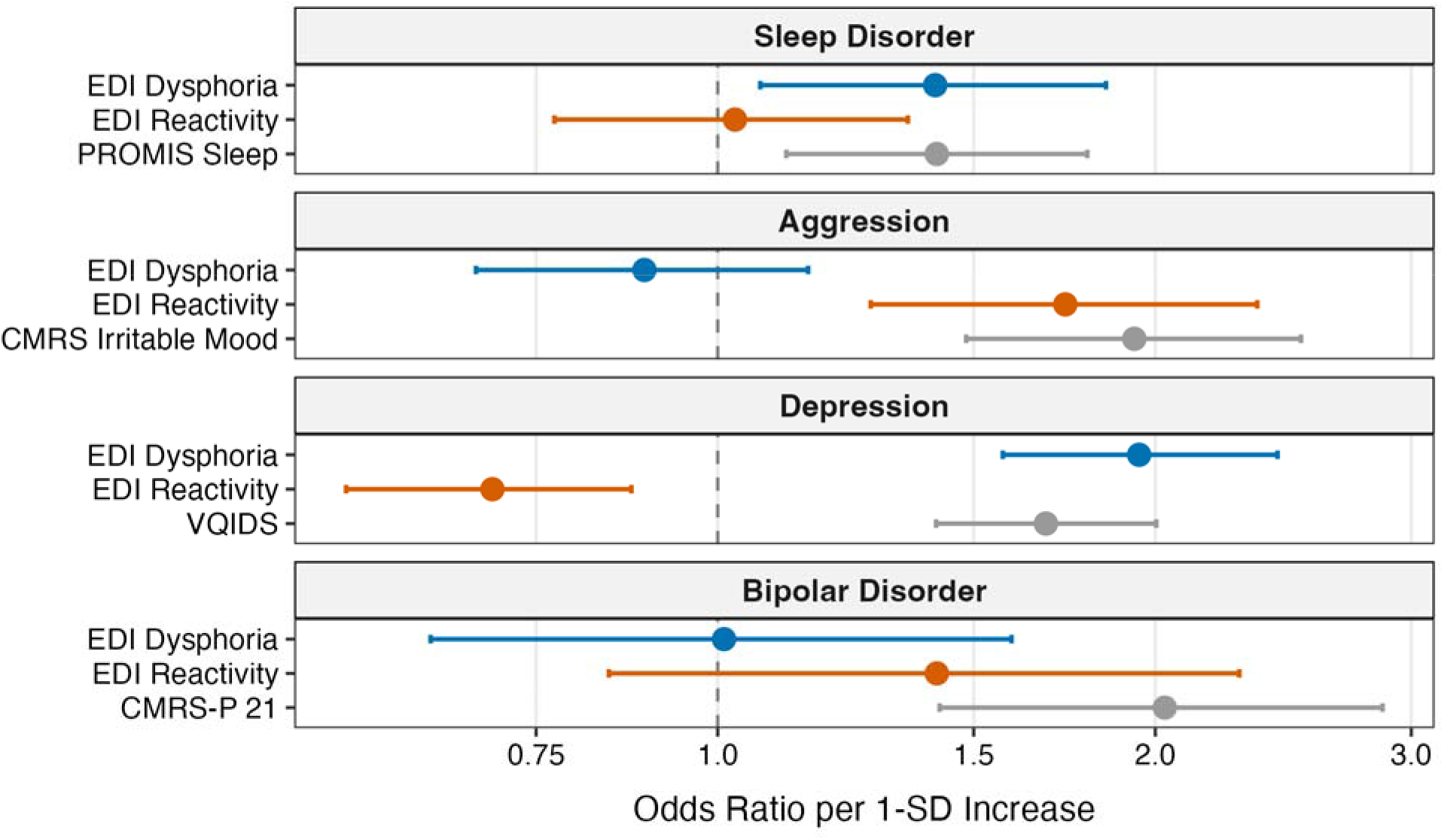
Odds ratios for different diagnoses associated with a 1 standard deviation increase in scale scores. ***Note.*** Points and horizontal bars represent adjusted odds ratios and 95% confidence intervals from separate logistic regression models. Each model included standardized EDI-Dysphoria and EDI-Reactivity scores together with the indicated domain-specific measure (PROMIS Sleep, CMRS-Irritable Mood, VQIDS, or CMRS-P 21). The dashed vertical line indicates an odds ratio of 1 (i.e., no difference). EDI-Dysphoria and EDI-Reactivity showed distinct conditional association patterns across outcomes.

## Discussion

The general factor accounted for 62% of common variance and was measured reliably, yet the provisional four-class solution distinguished a population-modal profile and three profiles characterized by internalizing symptoms, attention/hyperactivity symptoms, and unusual thought content with elevated mood. This addresses a practical question for evidence-based assessment, where scale selection often follows diagnostic clusters (Becker-Haimes et al., 2020; Hunsley & Mash, 2018; E. A. Youngstrom et al., 2020): whether a battery retains meaningful construct-specific information beyond general emotion dysregulation. Overall severity and symptom configuration appear to provide complementary perspectives on children’s difficulties. Evidence for these complementary perspectives is stronger, however, than evidence that four classes constitute a uniquely supported or reproducible classification.

The original raw-score analysis on a narrower battery found only a two-class severity gradient. Broadening the battery produced recognizable counterparts to the final profiles, but with diffuse elevations on unrelated indicators and unstable enumeration (an empty profile in every solution beyond two classes). Removing shared general-severity variance yielded more differentiated profiles with external correlates. This also informs scale selection: specific-factor indicators require reliable variance after general severity is partialled out, as assessed by the ω*_HS_*, *ECV*_SS_, and *H* benchmarks in Table 2.

That purification recovered a categorical triple dissociation, not just a severity ladder. ADHD-spectrum, mood/internalizing, and mania/thought-disorder-spectrum problems sorted into three classes with largely non-overlapping diagnostic, functional, and demographic signatures (supplemental Table S3) — evidence against a purely dimensional, undifferentiated view of pediatric transdiagnostic risk, at least at this level of resolution. This does not rest on model fit alone. The classes replicate on external, non-circular criteria: a consistent triple dissociation in caregiver-reported diagnoses and service use (attention/hyperactivity and developmental morbidity for Reactive/ADHD; depression, anxiety, and therapy use for Depressive/Internalizing; bipolar and sleep-disorder diagnoses for Thought/Mania); a demographic gradient matching known epidemiology (younger, more male, less pubertally advanced for Reactive/ADHD, older, more female, more pubertally advanced for Depressive/Internalizing) that held within each sex separately; and, on direct re-administration roughly two weeks later in an independent recontact subsample, real-world classification agreement well above chance (kappa=.60, Gwet’s AC1/AC2=.74) — figures that compare favorably to test-retest reliability for individual categorical DSM-5 diagnoses in the field trials (Regier et al., 2013), despite the harder four-way assignment task. An independent, from-scratch reproducibility audit — a full re-derivation of the pipeline against the raw data, not merely a re-run of cached results — reproduced essentially every one of these figures closely, including the reliability gate, the incremental-validity odds ratios to three decimal places, and the recontact kappa within a point.

CMRS-Irritable Mood’s lack of specific-factor signal directly replicates the central finding of the pediatric bipolar/DMDD nosology debate, rather than reflecting a psychometric weakness of this particular scale. Across every test in this study, CMRS-Irritable Mood’s *g*-purified specific factor carried essentially no independent signal: it was tied for second-weakest of the model’s 13 specific factors on the reliability gate (ω*_HS_*=.23, alongside EDI-Dysphoria), it correlated near zero or negatively with EDI-dysregulation, Vanderbilt-Hyperactivity/Inattention, and PSC-Externalizing when tested individually, and in the unified model it was negatively associated with all three ADHD-domain specific factors, most strongly with PSC-Attention (*r*=−.41). This is precisely what roughly three decades of nosology research converged on after considerable false starts: irritability, once separated from general symptom severity, does not specifically mark bipolar disorder, ADHD, or disruptive behavior — it behaves as a transdiagnostic feature, not a differentiating one (Leibenluft et al., 2024). Distinct types of aggressive behavior can still be measured and differentiated (Kahn et al., 2013; Langfus, Findling, Fatourou, et al., 2026; Langfus, Findling, Reynolds, et al., 2026), but irritability as a general construct is not itself diagnostically specific.

Luckenbaugh et al. (2009) similarly found that elevated mood and decreased need for sleep, rather than irritability, distinguished prospective symptom trajectories in pediatric bipolar disorder and ADHD. Sleep disturbance, episodic symptoms, and elated mood also feature in bipolar-onset risk calculators (Soehner et al., 2019; Van Meter et al., 2019). In our profiles, residual CMRS-Irritable Mood was mildly elevated in Depressive/Internalizing and, to a lesser extent, Thought/Mania, but suppressed in Reactive/ADHD. This pattern is consistent with DSM-5’s placement of DMDD among depressive disorders (American Psychiatric Association [APA], 2022), although limited irritability-specific reliability constrains its interpretation. The convergence with prior nosology research suggests caution in expecting irritability items to discriminate among presentations in a transdiagnostic mood battery, consistent with evidence of irritability’s cross-diagnostic associations in youth (Chin et al., 2025).

The recontact comparison should be read with the retest interval in mind. The roughly two-to-three-week gap between administrations is long enough that some Time-1/Time-2 disagreement plausibly reflects genuine short-term symptom change rather than pure classification unreliability, making the kappa a conservative, if anything, estimate of the model’s true short-term reliability. The pattern of disagreement across classes is itself informative: Reactive/ADHD showed the highest short-term recurrence of the three symptomatic classes (72.7%), consistent with ADHD’s neurodevelopmental, chronic course, while Depressive/Internalizing and Thought/Mania — the more episodic, state-dependent mood presentations — showed lower recurrence (53.3% and 57.1%, respectively). A short retest window should be expected to catch genuinely episodic phenomena mid-fluctuation more often than it catches a chronic, trait-like presentation shifting classes, so this pattern reads as evidence for, not against, the classes’ construct validity.

Although this analysis is theoretically similar to the Hierarchical Taxonomy of Psychopathology (HiTOP; Kotov et al., 2017, 2021) for understanding dimensional psychopathology, restricting the item pool to mood and related symptoms surfaced a structure that does not map cleanly onto HiTOP’s own spectra and subfactor levels. We found a potentially cross-cutting grouping of mood-related symptoms spanning both internalizing and externalizing content (dysphoria and reactivity together), alongside a combination of internalizing and somatoform (sleep) content that HiTOP conceptualizes under a general emotional-dysfunction grouping. Part of this difference may trace to the EDI itself: to our knowledge, no prior HiTOP-oriented study has used it to examine the structure of dimensional psychopathology, so this may be new evidence of how a transdiagnostic, cross-cutting mood measure interacts with the HiTOP hierarchy rather than a disagreement about the hierarchy itself.

The contrast with the earlier raw-score analysis suggests that the general-population sampling frame alone did not explain its lack of profile differentiation. Despite floor effects and modest class sizes, broadening the battery and separating general severity revealed differentiated profiles. Replication in clinically ascertained transdiagnostic samples, such as outpatient, school-based mental health, or partial hospital programs, would test generalizability at higher symptom levels and enrich representation of uncommon or more severe presentations (e.g., McClellan et al., under review). Similar profiles, with a smaller normative group, would support generalizability; additional profiles might emerge that were too rare or severe to be represented adequately here.

Caregiver-reported aggression/disruptive-behavior diagnoses were most elevated in the Reactive/ADHD profile, although smaller elevations were also observed in the Depressive/Internalizing and Thought/Mania profiles. This pattern is consistent with the frequent co-occurrence of emotional and behavioral difficulties, but it does not establish that one domain causes or explains the other. Prior work has linked depressive symptoms with reactive aggression and has documented reciprocal associations between internalizing and externalizing problems over time (Li et al., 2025; Yang et al., 2024). Longitudinal, multi-informant research is needed to clarify how broad symptom burden and domain-specific difficulties develop together.

Our findings map onto the prediction–prescription–process structure of evidence-based assessment (E. A. Youngstrom, Van Meter, et al., 2017). The general factor and the bifactor-purified profile shape are prediction-phase tools, not diagnoses. The general factor functions as an efficient, low-cost screen for overall risk — the kind of broad, high-sensitivity measure the evidence-based-assessment literature recommends for an initial, “vital few” pass (Youngstrom et al., 2017) — while the residual profile shape, net of that general severity, narrows a small set of competing hypotheses about which specific presentation is most likely and, correspondingly, where the more intensive prescription-phase work (structured interview, collateral report, domain-specific instruments) should concentrate. This is the same logic as the test zone between wait-test and test-treat thresholds in Bayesian clinical assessment (Youngstrom et al., 2017): a profile pointing toward a thought problems/manic group raises the local probability of bipolar disorder enough to warrant targeted follow-up without asserting the diagnosis outright, the same way a positive family history or an elevated CMRS score would shift probability in a clinical Bayesian workflow (Youngstrom, 2026).

The specific-factor reliability results carry a direct implication for what belongs in that follow-up assessment. PROMIS Pediatric Sleep-Related Impairment was the single most reliable and construct-replicable specific factor in the entire model (ω*_HS_*=.60, *H*=.92), and pubertal timing differentiated the Depressive/Internalizing and Reactive/ADHD classes independent of sex. Both findings converge with a direct clinical recommendation from the broader literature: sleep, pubertal development, and family psychiatric history and impact (the FIRM) function as supplemental assessments that help separate mood disorder from other sources of emotional and behavioral dysregulation, and belong in a standard pediatric assessment kit alongside brief mood scales (Youngstrom, 2026). Our data give that recommendation direct empirical support from a general-population sample: sleep-related impairment was not just clinically relevant but psychometrically the cleanest specific-factor signal in the entire battery, and pubertal timing tracked class membership even after accounting for sex.

PSC-Attention illustrates the other side of this same scale-selection logic. It cleared none of the three specific-factor reliability benchmarks (ω*_HS_*=.14, *ECV*_SS_=.19, *H*=.43) yet was retained as an explicit, disclosed exception because it has independently gained clinical traction as its own subscale. A companion sensitivity analysis found the retained four-class solution to be substantially dependent on this one weak indicator (adjusted Rand index=.43 when it is dropped alone, versus .92 when it is dropped together with CMRS-Irritable Mood) — a concrete illustration of the tension between psychometric guidance and clinical familiarity in scale selection, and a reason to treat PSC-Attention’s contribution to the profile solution with more caution than its face validity alone would suggest.

EDI-Dysphoria may provide the strongest broad screening signal for mood and behavioral difficulties, while residual phenotypic differences may warrant targeted assessment. Both EDI dimensions added selectively relevant information beyond domain-specific measures, with opposite associations with depression when modeled together. The negative Reactivity coefficient describes its association holding correlated predictors constant; it does not indicate protection against depression. Without partialling out a general factor, the two scales correlate highly and positively, consistent with other samples (Mazefsky, Conner, et al., 2021; Mazefsky, Day, et al., 2018). Thus, the general mood dimension may inform degree of difficulty, while specific dimensions help identify where further assessment should concentrate.

This transdiagnostic mood and behavioral battery offers several feasibility advantages consistent with this prediction-phase role. The 87-item battery can be completed in under 15 minutes, providing a strong utility-to-burden ratio given the breadth of clinical content covered, and every measure is publicly available at no cost with normative data available through the published literature or companion papers in this special issue. This battery is not intended to replace comprehensive, multimodal clinical evaluation, and associations among measures completed by the same caregiver cannot substitute for independent diagnostic assessment; rather, it is a pragmatic, cost-free prediction-phase tool for generating and prioritizing hypotheses that the next, more resource-intensive phase of assessment can then pursue.

## Limitations

Limitations of the current study include the lack of a known clinical group defined by best-practice clinical assessment or standardized structured clinical interviews, and the presence of significant floor effects across all included measures. Although we found some evidence of divergent validity of the general mood profile, we did not include direct measures of other clinical conditions (e.g., autistic spectrum, ADHD, anxiety), and all assessments were caregiver (proxy) report rather than multi-informant or multimodal. The design was cross-sectional, which precludes claims about developmental stability or transition between profiles over time.

Additionally, the data were all proxy report. The bipolar-disorder criterion rested on a small number of caregiver-reported cases and warrants replication with a larger or clinically-enriched sample, and the four-class latent profile solution showed modest, not perfect, multi-start agreement (adjusted Rand index .69–.91). A post-hoc reproducibility audit additionally found the retained solution to be sensitive to a single specific factor: omitting PSC-Attention alone from the profile indicators and refitting yielded only fair agreement with the retained solution (adjusted Rand index=.43), whereas omitting both PSC-Attention and CMRS-Irritable Mood together yielded much closer agreement (adjusted Rand index=.92); because PSC-Attention is also the one specific factor that failed every reliability benchmark in Table 2, it may be doing more work in shaping the retained class structure than its psychometric profile alone would suggest. Finally, the general-population sampling frame likely limits statistical power for the rarest phenotype-diagnosis pairings. Additional research incorporating multimodal, multi-informant assessment methods, particularly within clinical and longitudinal samples, is needed to extend and replicate these results.

## Clinical implications

These findings do not support using profile membership as a diagnosis or stand-alone basis for treatment decisions. In this general-population sample, however, they suggest value in distinguishing shared symptom burden from relative symptom patterns across domains. Sleep-related impairment, depressive symptoms, attention/hyperactivity symptoms, and unusual-thought/elevated-mood content contributed differently across retained profiles, whereas irritability showed limited differentiation after accounting for shared variance.

Incremental-validity analyses suggest that EDI domains may complement established symptom measures. EDI-Dysphoria was associated with caregiver-reported depression beyond the brief depression measure, whereas EDI-Reactivity was associated with caregiver-reported aggression/disruptive behavior beyond the CMRS-P Irritable Mood scale. This pattern supports distinguishing Dysphoria from Reactivity rather than treating them as interchangeable indicators of general emotional distress.

Broad assessment may also help contextualize specific symptom elevations. Broadband measures commonly yield substantially overlapping total and syndrome scores. A bifactor-based scoring framework could supplement established measure-specific scores by distinguishing overall symptom burden from domains that are relatively prominent or diminished. For example, a youth may have elevated norm-referenced scores for sleep impairment, depression, and attention problems. A high *g* score without a sleep-specific elevation would suggest that sleep difficulties reflect broadly elevated symptoms; a high sleep-specific factor score would suggest disproportionate sleep impairment relative to overall burden. Established measure scores would indicate absolute elevation, while bifactor scores would contextualize the pattern given the youth’s overall psychological distress. When difficulties span many domains, this approach could help identify priorities for further assessment and inform treatment planning alongside caregiver input and preferences. Any bifactor-based scoring system would require independent validation before clinical use.

## Data Availability

All data produced in the present study are available upon reasonable request to the authors.

## Acknowledgements

Supported by gifts from the Chlapaty, DiMarco, and Zipfel families to Nationwide Children’s Hospital to support the Institute for Mental and Behavioral Health Research (IMBHR). The authors gratefully acknowledge Jonathan Covarrubias for his assistance with the clinical dataset, and Marissa McClellan, Kevin Stephenson, Jacqui Pazaropoulos, Brittany Grate, and Kim Jones for their contributions to the development and implementation of the BENCHMARK project, including literature and reference review, study coordination and administration, data organization and management, data cleaning, and development of analytic resources.

## Disclosures

Dr. Eric A. Youngstrom is the co-founder and Executive Director of Helping Give Away Psychological Science, a 501c3; receives grant support from the National Institute of Health; he has received royalties from the American Psychological Association and Guilford Press, and he holds equity in Joe Startup Technologies.

## Notes

### Author Declarations

IRB of Nationwide Children's Hospital waived ethical approval for this work.

