## Supplemental files for "Parsing the Pediatric Mood Spectrum in U.S. Youth: Transdiagnostic Severity Versus Phenotypic Specificity"

**Supplemental Materials**

Figure S1. Schematic of the bifactor model fit to the item level data.

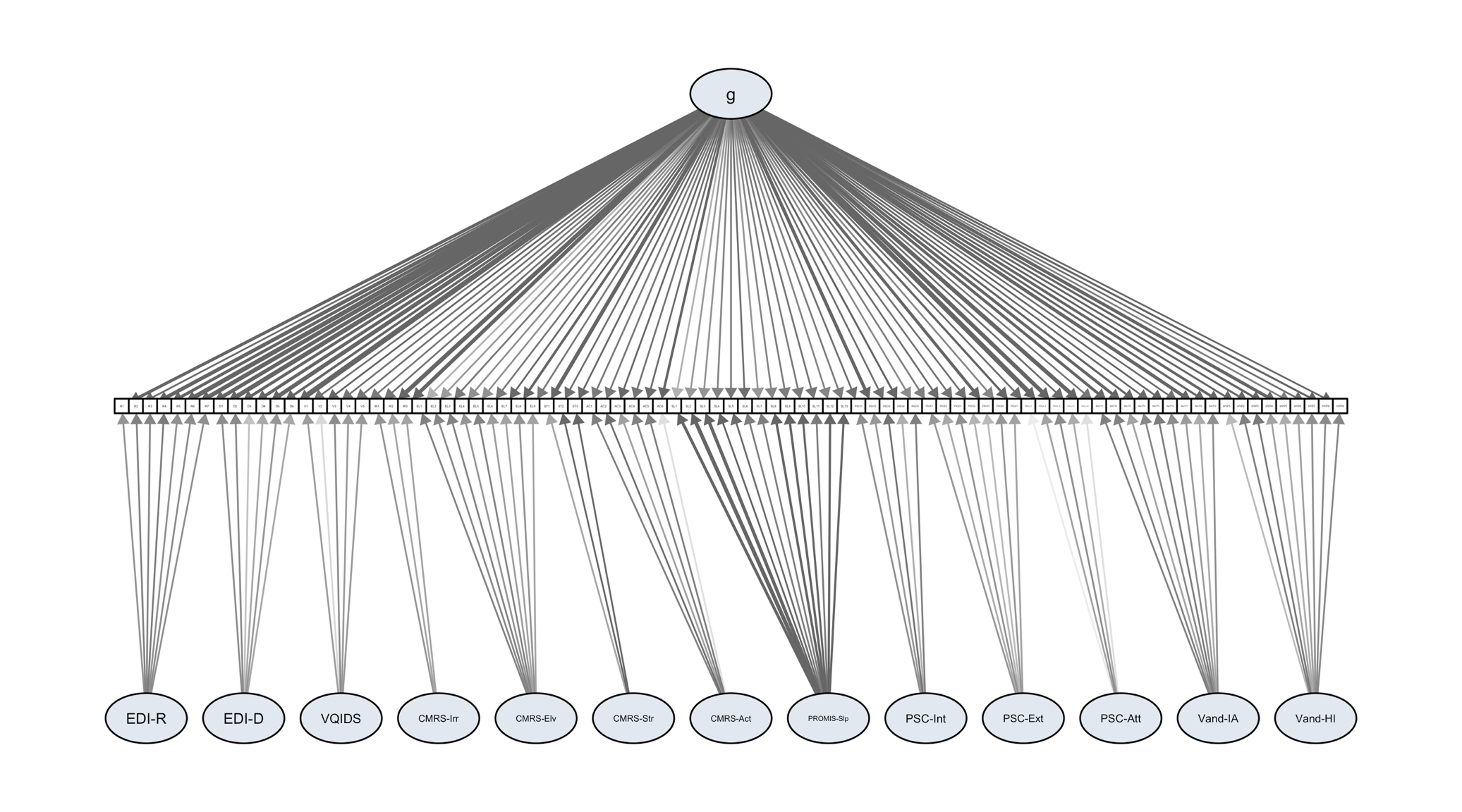

### Figure S2. Bifactor Measurement Model Structure

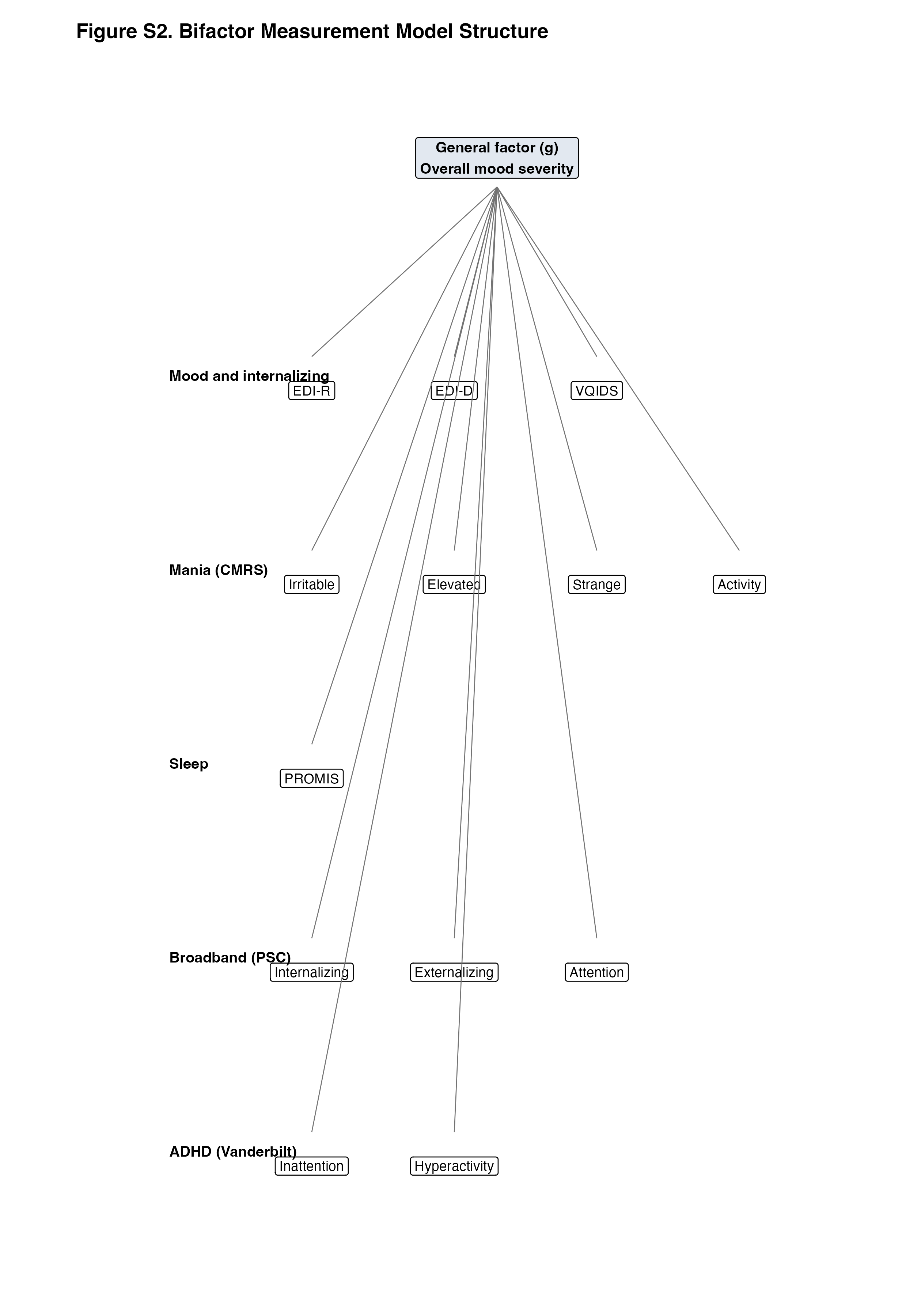

### Table S1. Class Centroids on the 14 Bifactor-Purified Indicators (*z*-scored *M* [*SD*])

| **Indicator** | Normative (*n*=1473, 67.4%) | Depressive/Internalizing (*n*=259, 11.8%) | Reactive/ADHD (*n*=240, 11.0%) | Thought/Mania (*n*=215, 9.8%) |
| --- | --- | --- | --- | --- |
| General factor (*g*) | -0.41 (0.78) | 0.66 (0.81) | 1.06 (0.78) | 0.82 (0.97) |
| EDI-Reactivity | 0.03 (0.91) | 0.07 (1.20) | -0.02 (1.24) | -0.26 (0.99) |
| EDI-Dysphoria | -0.05 (0.81) | 1.07 (0.94) | -1.03 (0.82) | 0.18 (1.13) |
| VQIDS (depression) | -0.20 (0.73) | 1.48 (0.84) | -0.57 (0.84) | 0.19 (1.25) |
| CMRS-Irritable | 0.00 (0.92) | 0.18 (1.07) | -0.55 (0.96) | 0.41 (1.22) |
| CMRS-Elevated | -0.01 (0.92) | -0.44 (0.86) | -0.26 (0.94) | 0.93 (1.15) |
| CMRS-Strange Thoughts | -0.25 (0.23) | -0.30 (0.57) | -0.51 (0.60) | 2.65 (1.08) |
| CMRS-Activity | -0.02 (0.92) | -0.47 (1.00) | 0.22 (1.22) | 0.49 (0.98) |
| PROMIS-Sleep | 0.09 (0.88) | 0.49 (0.95) | -1.22 (0.82) | 0.14 (1.00) |
| PSC-Internalizing | -0.09 (0.85) | 1.13 (0.97) | -0.57 (0.99) | -0.11 (1.02) |
| PSC-Externalizing | 0.10 (0.94) | -0.39 (1.05) | -0.02 (1.12) | -0.20 (1.05) |
| PSC-Attention | 0.00 (0.85) | -0.31 (1.01) | 0.96 (0.92) | -0.71 (1.19) |
| Vanderbilt-Inattention | -0.03 (0.89) | -0.13 (1.08) | 0.86 (1.03) | -0.59 (1.01) |
| Vanderbilt-Hyperactivity | 0.03 (0.86) | -0.77 (0.94) | 0.99 (1.00) | -0.38 (0.96) |

### Table S2. BCH-Weighted Demographic Estimates by Class

| **Variable** | **Normative** | **Depressive/Internalizing** | **Reactive/ADHD** | **Thought/Mania** |
| --- | --- | --- | --- | --- |
| Child age, years (6 to 18) | 11.65 | 13.60**** | 9.63**** | 12.33* |
| % male (binary) | 51.51 | 39.26** | 73.79**** | 50.64 |
| Family income  (bracket 1-16) | 8.16 | 7.16** | 7.40* | 7.35** |
| Pubertal development  (PDS total, 5-20) | 11.76 | 14.39**** | 8.63**** | 12.91*** |

***Note.*** Cells show BCH-weighted estimated means; % male shows the BCH-weighted estimated proportion male × 100. Stars denote cluster-robust BCH-weighted significance of the difference from Normative: **p* < .05, ***p* < .005, ****p* < .0005, *****p* < .00005, all two-tailed.

### Table S3. Diagnostic and Service-Use Criterion Validity by Class (BCH-weighted, sorted by Cramér's *V*)

| **Criterion** | **Cramér's *V*** | **Normative** | **Depressive/Internalizing** | **Reactive/ADHD** | **Thought/Mania** |
| --- | --- | --- | --- | --- | --- |
| Medication | .332 | 7.4% | +22.8%* | +37.9%**** | +29.0%**** |
| ADHD | .300 | 12.4% | +14.2%*** | +49.8%**** | +11.1%*** |
| Therapy | .294 | 18.3% | +46.0%**** | +40.6%**** | +12.5%* |
| Depression | .274 | 2.6% | +25.4%**** | +5.1%* | +18.5%**** |
| Anxiety | .246 | 7.9% | +32.7%**** | +17.5%**** | +15.3%**** |
| Autism | .211 | 3.8% | +8.0%** | +23.0%**** | +3.5% |
| OTC/supplement | .196 | 15.9% | -3.8% | +23.9%**** | +15.6%** |
| Aggression | .183 | 0.7% | +4.4%** | +12.9%**** | +5.3%** |
| Bipolar | .151 | 0.1% | +1.6% | +2.3%* | +5.6%*** |
| LD | .114 | 5.4% | +3.0% | +11.3%*** | +6.3%** |
| Sleep Disorder | .095 | 2.7% | +4.0%* | +3.6% | +6.2%** |

***Note.*** Normative column is the BCH-weighted estimated base rate; other columns are BCH-weighted percentage-point differences from Normative. Stars denote cluster-robust weighted-regression contrast significance: **p* < .05, ***p* < .005, ****p* < .0005, *****p* < .00005, all two-tailed. Cramér's *V* values are descriptive uncorrected effect sizes used for ordering the criteria.

### Table S4. Continuous Functional/Severity Validators by Class (BCH-weighted difference from Normative)

| **Validator** | **Potential Range** | **Normative** | **Depressive/Internalizing** | **Reactive/ADHD** | **Thought/Mania** |
| --- | --- | --- | --- | --- | --- |
| NQLS (quality of life) | 0ꜝ to 28 | 22.31 | -5.63**** | -4.86**** | -3.14**** |
| N-Gauge (global severity) | 0 to 5ꜝ | 0.33 | +1.79**** | +1.06**** | +0.85**** |
| FIRM-22 (family-history risk for mood disorders) | 0 to 22ꜝ | 2.47 | +2.61**** | +1.97**** | +1.49**** |
| CHRT (suicidality risk) | 0 to 56ꜝ | 4.67 | +11.65**** | +6.04**** | +10.22**** |
| Vanderbilt academic impairment | 4 to 20ꜝ | 8.24 | +1.94**** | +4.31**** | +1.04*** |
| Vanderbilt social impairment | 4 to 20ꜝ | 7.89 | +3.64**** | +3.75**** | +1.34**** |

ꜝIndicates the most impaired score possible. **p* < .05, ***p* < .005, ****p* < .0005, *****p* < .00005, all two-tailed.

### Table S5. Time 1 vs. Time 2 Class Assignment, Recontact Sample (N = 166)

| **Time 1 class** | **Depressive/ Internalizing** | **Reactive/ ADHD** | **Normative** | **Thought/ Mania** | **Row Total** | **% Same Class at  Time 2** |
| --- | --- | --- | --- | --- | --- | --- |
| Depressive/Internalizing | **16** | 0 | 9 | 5 | 30 | 53.3% |
| Reactive/ADHD | 0 | **16** | 6 | 0 | 22 | 72.7% |
| Normative | 3 | 4 | **95** | 5 | 107 | 88.8% |
| Thought/Mania | 2 | 0 | 1 | **4** | 7 | 57.1% |

***Note.*** Rows are Time-1 (original) class assignment; columns are Time-2 (recontact, ~2 weeks later) class assignment, scored against the fixed Time-1 bifactor + G=4 model rather than a refit model. Raw agreement = 78.9%; Cohen's kappa = .600 [.487, .713]; Gwet's AC1 = AC2 = .744 [.666, .823] (AC1 and AC2 coincide because these four classes are unordered nominal phenotypes with no ordinal structure to weight)*.*

### Table S6. Theoretical and Observed Score Ranges for Bifactor Model Indicators and Selected Validators

| **Scale** | **Items** | **Per-Item Range** | **Theoretical Range** | **Observed Range** |
| --- | --- | --- | --- | --- |
| **Bifactor Model Indicators (13 Specific Factors, 87 Items)** | | | | |
| EDI-Reactivity | 7 | 0–4 | 0–28 | 0–28 |
| EDI-Dysphoria | 6 | 0–4 | 0–24 | 0–24 |
| VQIDS | 5 | 0–3 | 0–15 | 0–15 |
| CMRS-Irritable Mood | 3 | 0–3 | 0–9 | 0–9 |
| CMRS-Elevated Mood | 9 | 0–3 | 0–27 | 0–23 |
| CMRS-Strange Thoughts | 3 | 0–3 | 0–9 | 0–9 |
| CMRS-Increased Activity | 6 | 0–3 | 0–18 | 0–18 |
| PROMIS Pediatric Sleep-Related Impairment | 13 | 1–5 | 13–65 | 13–65 |
| PSC-Internalizing | 5 | 0–2 | 0–10 | 0–10 |
| PSC-Externalizing | 7 | 0–2 | 0–14 | 0–14 |
| PSC-Attention | 5 | 0–2 | 0–10 | 0–10 |
| Vanderbilt-Inattention | 9 | 0–3 | 0–27 | 0–27 |
| Vanderbilt-Hyperactivity/Impulsivity | 9 | 0–3 | 0–27 | 0–27 |
| **Validators (Not Bifactor Model Indicators)** | | | | |
| FIRM (22-item) | 22 | 0–1 | 0–22 | 0–19 |
| NQLS | 7 | 0–4 | 0–28 | 0–28 |
| *Note. Analytic N = 2,187 for all rows except NQLS (N = 2,186; one case missing). Observed range is the range of summed scores actually attained in the analytic sample; theoretical range is the minimum and maximum attainable score given the number of items and per-item response range. Every scale reaches both the theoretical floor and ceiling except CMRS-Elevated Mood and FIRM, which do not reach the theoretical ceiling in this sample.* | | | | |
